# The Efficacy and Safety of Sodium-Glucose Cotransporter Protein 2 Inhibitors (SGLT2Is) in Liver Cirrhosis: A Systematic Review and Meta-Analysis

**DOI:** 10.1101/2024.12.27.24319638

**Authors:** Sudheer Dhoop, Mohammed Shehada, Bisher Sawaf, Manthan Patel, Luke Roberts, Wade-Lee Smith, Benjamin Hart

## Abstract

**Background:** Sodium-Glucose Cotransporter Protein 2 Inhibitors (SGLT2Is) are known for their cardiovascular and renoprotective benefits and are efficacious in managing Metabolic-Associated Steatotic Liver Disease (MASLD). However, limited data exist on their use in advanced liver disease, particularly liver cirrhosis.

**Aims:** To synthesize existing evidence on the efficacy and safety of SGLT2Is in patients with liver cirrhosis and to provide clinical guidance.

**Methods:** A systematic review and meta-analysis were conducted following the PRISMA 2020 Statement. Searches in major health databases identified studies where SGLT2Is were used in patients with cirrhosis. The analysis focused on prospective trials in decompensated cirrhosis and retrospective studies in compensated cirrhosis. Primary outcomes included the need for large-volume paracentesis (LVP) and mortality. Secondary outcomes assessed weight loss, loop diuretic dose reduction, residual ascites, acute kidney injury (AKI), hyponatremia, hepatic encephalopathy (HE), and urinary tract infections (UTIs).

**Results:** Ten studies (8 peer-reviewed) from 2020-2024 were included: 2 randomized controlled trials, 4 single-arm prospective trials, and 4 retrospective studies. SGLT2I use was associated with reduced LVP (RR 0.45, CI 0.31-0.66, p<0.001) and mortality (aHR 0.46, CI 0.38-0.55, p<0.001). Benefits included a 39 mg reduction in loop diuretic dose, 7 kg weight loss, and no significant increase in residual ascites, AKI, hyponatremia, HE, or UTIs.

**Conclusions:** SGLT2Is show promise in managing diuretic-resistant ascites and reducing mortality in liver cirrhosis without causing significant adverse events. Larger, randomized controlled trials are needed to validate these findings.

## Introduction

Sodium-glucose cotransporter protein 2 inhibitors (SGLT2Is) were initially introduced as therapeutic agents for type 2 diabetes mellitus, primarily by inhibiting sodium-glucose cotransporters in the proximal renal tubule, thereby increasing glucosuria and improving glycaemic control [1]. Over time, a growing body of evidence has highlighted their cardiovascular advantages [2] and reno-protective properties [3], benefits that may partly stem from their modest diuretic effect [1]. Although substantial literature supports the use of SGLT2Is in the early stages of metabolic dysfunction-associated steatotic liver disease (MASLD) [4–7], including their ability to reduce liver stiffness, lower hepatic enzyme levels, decrease visceral adiposity, improve insulin sensitivity, and potentially prevent the progression to cirrhosis, data on their role in advanced liver disease remain sparse [8].

Cirrhosis, a late-stage manifestation of chronic liver injury, is a clinical diagnosis defined by histopathological findings of bridging fibrosis [9] and is characterised clinically by impaired hepatic synthetic capacity, metabolic dysfunction, and development of portal hypertension [10]. Cirrhosis follows a clinical course that may be initially compensated, with few overt clinical signs [11], but can evolve into decompensated disease marked by complications such as large-volume ascites, variceal bleeding secondary to portal hypertension [12], hepatic encephalopathy [13], and hepatorenal syndrome (HRS) due to severe circulatory and metabolic derangements [14].

Within this context, there is growing interest in leveraging the multifaceted benefits of SGLT2Is in cirrhosis. These agents may slow progression of cirrhosis secondary to MASLD through their cardiovascular [2] and anti-fibrotic benefits [6,7], as well as possibly reducing portal hypertension through their diuretic action, as suggested by preclinical animal models [15].

Nonetheless, clinical caution is warranted. The glycosuric effect that facilitates SGLT2Is’ therapeutic benefit in diabetes also predisposes patients to genitourinary infections, which can be more severe in patients with cirrhosis due to compromised immune function [16]. In addition, as with loop diuretics [17], the haemodynamic instability inherent to cirrhotic patients—who often have baseline hypotension [10] and renin-angiotensin-aldosterone system (RAAS) dysregulation—can amplify the risk of acute kidney injury (AKI) and/or HRS [14], affect electrolyte homeostasis [8], potentially worsen hepatic encephalopathy [18], and possibly contribute to sarcopenia [19], which negatively influences long-term outcomes in cirrhosis [20]. Many retrospective database studies [21–24] have demonstrated that SGLT2I use may be associated with reduced mortality and hepatic decompensation events in patients with cirrhosis, with data suggesting benefit not only in compensated disease [20–23] but also in decompensated states [24]. Likewise, case reports [25,26] and case series [27], as reviewed by several systematic and narrative syntheses [4,28–30], have outlined scenarios in which SGLT2Is, despite their potential adverse effects, may serve as valuable adjunctive treatments for managing refractory ascites when combined with conventional diuretics such as loop diuretics and mineralocorticoid receptor antagonists [17,31]. In recent years, a series of experimental trials and additional case series have broadened this evidence base, focusing on the role of SGLT2Is in cirrhosis-related fluid management [32–38].

Still, formal guidelines currently advise against routine SGLT2I use in cirrhotic populations due to insufficient evidence and safety concerns [30]. Therefore, we aim to provide a comprehensive systematic review and meta-analysis of studies examining both the short- and long-term efficacy and safety of SGLT2Is in cirrhotic patients. Our objectives include evaluating their utility as diuretic adjuncts, delineating their adverse event profile, and assessing their potential to confer meaningful long-term benefits. Synthesis of results will include meta-analysis, but also critical appraisal of each outcome, accounting for the limitations and differences of each study, culminating in a comprehensive review of SGLT2I use in liver cirrhosis to assist clinicians faced with complex decision-making in this high-risk population.

## Methods

### Search Strategy

A comprehensive search strategy to identify studies involving patients with cirrhosis exposed to SGLT2Is was developed in Embase (Embase.com, Elsevier) by an experienced health sciences librarian (WL-S) using truncated keywords, phrases, and subject headings (see Supplementary Table 1). This strategy was translated to MEDLINE (PubMed platform, National Center for Biotechnology Information, National Library of Medicine), Cochrane Central Register of Controlled Trials (CochraneLibrary.com, Wiley), Web of Science Core Collection, Korean Citation Index (Web of Science platform, Clarivate), and Global Index Medicus (World Health Organization). The initial search was conducted on 28 October 2024 (Supplementary Table 2). No publication date or language limits were applied. All results were exported to EndNote 21 citation management software (Clarivate, Philadelphia, PA, USA).

### Study Screening

To screen studies, duplicates were removed through successive iterations of EndNote’s duplicate detection algorithms and manual inspection. Two authors (S.D. and B.S.) reviewed the records and excluded duplicates not removed by the software, studies involving animals or children, review articles, case series, and study protocols. Experimental and observational studies published in all languages and countries were included. Studies were excluded if SGLT2Is were not the intervention or if the entire study population did not have cirrhosis. Discrepancies in study inclusion or exclusion were resolved by an independent reviewer (M.P.). The study selection process followed the flow diagram recommended by the PRISMA 2020 statement [39] (see Figure 1).

**Figure 1.).**
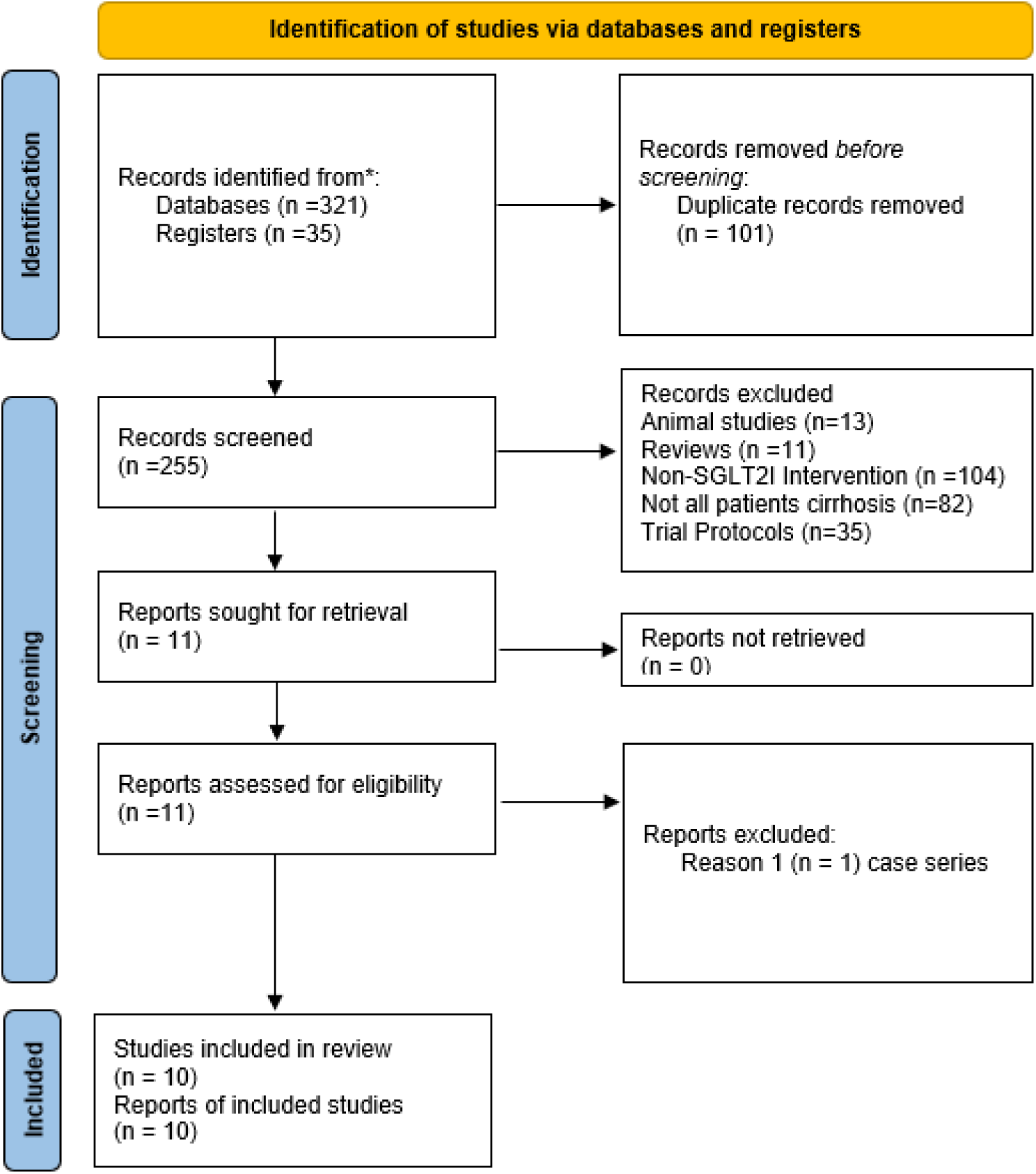
PRISMA 2020 flow diagram for new systematic reviews which included searches of databases and registers only

### Extraction of Raw Data and Outcome Selection

All studies that met the inclusion criteria were reviewed. The following data were extracted and summarised in Table 1: study design, data source, controlled or propensity-matched variables, sample size, percentage of SGLT2I distribution, percentage with decompensated cirrhosis, percentage with non-alcoholic steatohepatitis (NASH) cirrhosis, study period, and efficacy and safety outcomes. Data extraction was performed by S.D., cross-checked by B.S., and recorded in a Microsoft Excel spreadsheet. Table 2 summarises the reported outcomes.

**Table 1.**
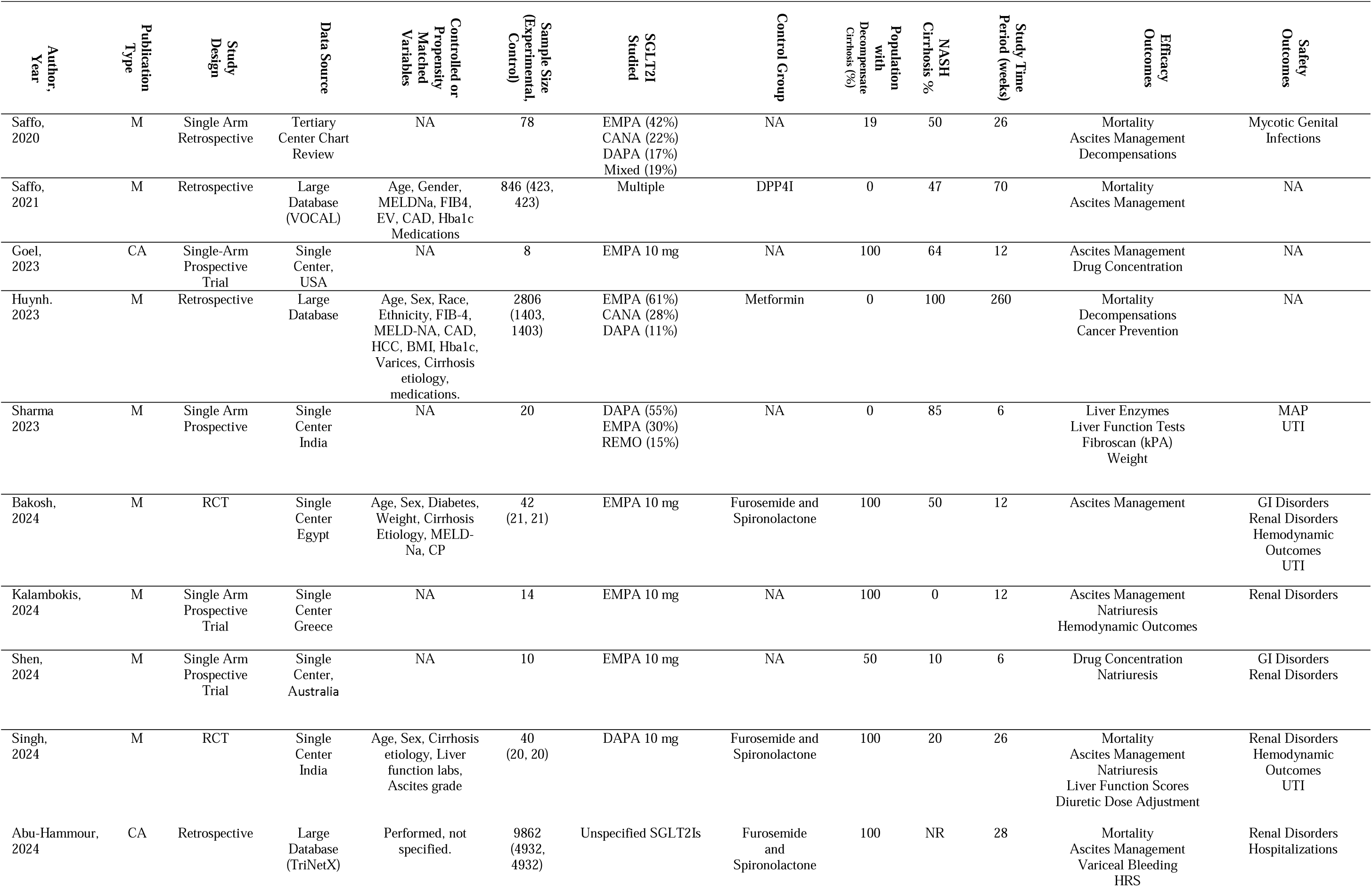
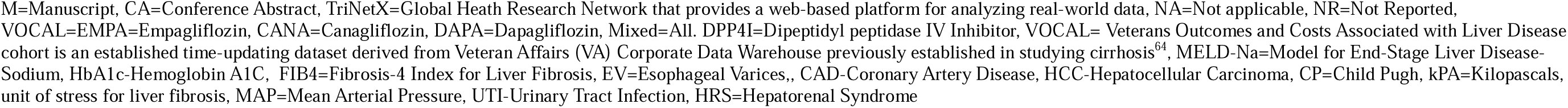
Characteristics for studies assessing SGLT2I as an intervention in liver cirrhosis.

| Author, Year | Publication Type | Study Design | Data Source | Controlled or Propensity Matched Variables | Sample Size (Experimental Control) | SGLT2I Studied | Control Group | Population with Decompensate Cirrhosis (%) | NASH Cirrhosis % | Study Time Period (weeks) | Efficacy Outcomes | Safety Outcomes |
| --- | --- | --- | --- | --- | --- | --- | --- | --- | --- | --- | --- | --- |
| Saffo, 2020 | M | Single Arm Retrospective | Tertiary Center Chart Review | NA | 78 | EMPA (42%)<br>CANA (22%)<br>DAPA (17%)<br>Mixed (19%) | NA | 19 | 50 | 26 | Mortality<br>Ascites Management<br>Decompensations | Mycotic Genital Infections |
| Saffo, 2021 | M | Retrospective | Large Database (VOCAL) | Age, Gender, MELDNa, FIB4, EV, CAD, HbA1c Medications | 846 (423, 423) | Multiple | DPP4I | 0 | 47 | 70 | Mortality<br>Ascites Management | NA |
| Goel, 2023 | CA | Single-Arm Prospective Trial | Single Center, USA | NA | 8 | EMPA 10 mg | NA | 100 | 64 | 12 | Ascites Management<br>Drug Concentration | NA |
| Huynh, 2023 | M | Retrospective | Large Database | Age, Sex, Race, Ethnicity, FIB-4, MELD-NA, CAD, HCC, BMI, HbA1c, Varices, Cirrhosis etiology, medications. | 2806 (1403, 1403) | EMPA (61%)<br>CANA (28%)<br>DAPA (11%) | Metformin | 0 | 100 | 260 | Mortality<br>Decompensations<br>Cancer Prevention | NA |
| Sharma 2023 | M | Single Arm Prospective | Single Center India | NA | 20 | DAPA (55%)<br>EMPA (30%)<br>REMO (15%) | NA | 0 | 85 | 6 | Liver Enzymes<br>Liver Function Tests<br>Fibroscan (kPA)<br>Weight | MAP<br>UTI |
| Bakosh, 2024 | M | RCT | Single Center Egypt | Age, Sex, Diabetes, Weight, Cirrhosis Etiology, MELD-Na, CP | 42 (21, 21) | EMPA 10 mg | Furosemide and Spironolactone | 100 | 50 | 12 | Ascites Management | GI Disorders<br>Renal Disorders<br>Hemodynamic Outcomes<br>UTI |
| Kalambokis, 2024 | M | Single Arm Prospective Trial | Single Center Greece | NA | 14 | EMPA 10 mg | NA | 100 | 0 | 12 | Ascites Management<br>Natriuresis<br>Hemodynamic Outcomes | Renal Disorders |
| Shen, 2024 | M | Single Arm Prospective Trial | Single Center, Australia | NA | 10 | EMPA 10 mg | NA | 50 | 10 | 6 | Drug Concentration<br>Natriuresis | GI Disorders<br>Renal Disorders |
| Singh, 2024 | M | RCT | Single Center India | Age, Sex, Cirrhosis etiology, Liver function labs, Ascites grade | 40 (20, 20) | DAPA 10 mg | Furosemide and Spironolactone | 100 | 20 | 26 | Mortality<br>Ascites Management<br>Natriuresis<br>Liver Function Scores<br>Diuretic Dose Adjustment | Renal Disorders<br>Hemodynamic Outcomes<br>UTI |
| Abu-Hammour, 2024 | CA | Retrospective | Large Database (TriNetX) | Performed, not specified. | 9862 (4932, 4932) | Unspecified SGLT2Is | Furosemide and Spironolactone | 100 | NR | 28 | Mortality<br>Ascites Management<br>Variceal Bleeding<br>HRS | Renal Disorders<br>Hospitalizations |

M=Manuscript, CA=Conference Abstract, TriNetX=Global Health Research Network that provides a web-based platform for analyzing real-world data, NA=Not applicable, NR=Not Reported, VOCAL=EMPA=Empagliflozin, CANA=Canagliflozin, DAPA=Dapagliflozin, Mixed=All. DPP4I=Dipeptidyl peptidase IV Inhibitor, VOCAL= Veterans Outcomes and Costs Associated with Liver Disease cohort is an established time-updating dataset derived from Veteran Affairs (VA) Corporate Data Warehouse previously established in studying cirrhosis<sup>64</sup>, MELD-Na=Model for End-Stage Liver Disease-Sodium, HbA1c-Hemoglobin A1C, FIB4=Fibrosis-4 Index for Liver Fibrosis, EV=Esophageal Varices., CAD-Coronary Artery Disease, HCC-Hepatocellular Carcinoma, CP=Child Pugh, kPA=Kilopascals, unit of stress for liver fibrosis, MAP=Mean Arterial Pressure, UTI-Urinary Tract Infection, HRS=Hepatorenal Syndrome

**Table 2A:** Outcomes in Prospective Trials.

| Author,<br>Year | Intervention | Residual<br>Ascites | Need for<br>LVP | Acute<br>Kidney<br>Injury | Hyponatremia | Hepatic<br>Encephalopathy | Urinary Tract<br>Infections | Diuretic Dose<br>Reduction (mg) | Weight Reduction<br>(kg) and 95% CI |
| --- | --- | --- | --- | --- | --- | --- | --- | --- | --- |
|  | Control (if applicable) |  |  |  |  |  |  |  |  |
| Goel,<br>2023 | Empagliflozin | NR | NR | 0/8 | NR | 0/8 | NR | 20 | -16.7 (+4.2, -37.6) |
| Sharma<br>2023 | SGLT2I | NR | NR | NR | NR | NR | NR | NR | -3.8 (+1.83,-8.63) |
| Bakosh,<br>2024 | Empagliflozin plus MRA+<br>Furosemide | 16/21 | 9/21 | 7/21 | 9/21 | 9/21 | 7/21 | NR | -7.0 (-3.6, -10.4) |
|  | MRA+<br>Furosemide | 21/21 | 21/21 | 12/21 | 2/21 | 6/21 | 2/21 | NR | NA |
| Kalambokis, 2024 | Empagliflozin | 7/14 | 0/14 | 0/14 | NR | NR | 0/14 | 140±9 to 77±11 | -7.2 (-2.8, -11.5) |
| Shen,<br>2024<br>(Decompensated) | Empagliflozin | NR | NR | NR | NR | NR | 0/5 | NR | NR |
| Singh,<br>2024 | Dapagliflozin plus MRA+<br>Torsemide | 17/20 | 7/20 | 10/20 | 5/20 | 2/20 | 11/20 | 9.4 ± 1.9 (188+/-38)<br>to 6.1 ± 4.3 (122+/-<br>86) | NR |
|  | MRA+ Torsemide | 20/20 | 15/20 | 3/20 | 3/20 | 1/10 | 4/20 | 9.2 ± 17 mg to 9 ±<br>2.5 mg | NR |

**Table 2B:**
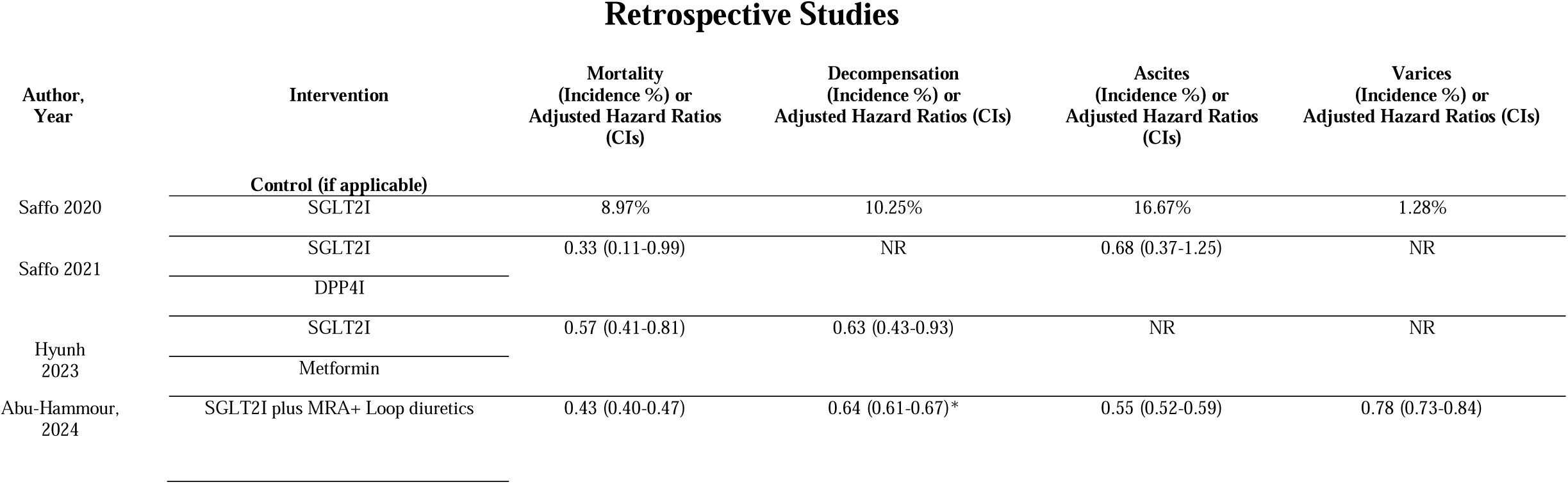
Outcomes for Retrospective Studies.

For prospective trials, the incidence of requiring large-volume paracentesis (LVP) was extracted as the primary dichotomous outcome, as diuretics are used to prevent the need for further ascites intervention [10]. Secondary dichotomous outcomes included the incidence of residual ascites, acute kidney injury (AKI), hyponatraemia, hepatic encephalopathy (HE), and urinary tract infections (UTIs). Continuous outcomes, such as weight reduction (in kilograms) and loop diuretic dose reduction (in milligrams), were also recorded. For randomised controlled trials, data were extracted from the experimental group for single-arm meta-analysis [40]. All outcomes were measured at study endpoints.

For retrospective studies, adjusted hazard ratios (aHR) with confidence intervals (CIs) for mortality were extracted as the primary outcome. Secondary outcomes included aHRs for hepatic decompensations (defined as a composite of variceal bleeding, HE, and ascites), ascites alone, and varices alone.

### Data Standardization

Before meta-analysis, weight and diuretic dose reduction data were standardised. For weight reduction, all data were converted to the mean, confidence interval, and sample size. One single-arm study [34] reported weight loss in litres removed with a *p*-value. Kilograms were approximated using a 1:1 ratio based on ascitic fluid having a density similar to water [41]. Confidence intervals were derived using inverse probability transformation of the *p*-value [42]. Another single-arm study expressed weight reduction as baseline and post-intervention means ± standard error. The standard error of the difference formula for two independent means [42] was used to calculate the mean and confidence intervals (Table 2).

For diuretic dose, two studies were pooled: one used furosemide [32] and the other torsemide [33]. A conversion factor of 2.5:1 was applied [44]. For retrospective studies, aHRs for varices and ascites were pooled using inverse variance weighting [42] to calculate an overall aHR for hepatic decompensation.

### Meta-Analysis

Statistical analysis was conducted using Comprehensive Meta-Analysis (CMA) Version 4.0. Prospective and retrospective trials were pooled separately due to their distinct outcomes. For prospective studies, dichotomous outcomes (residual ascites, need for LVP, AKI, HE, and UTI) were pooled to generate risk ratios (RR) with confidence intervals using a random-effects model. Continuous outcomes (loop diuretic dose and weight reduction) were also pooled using a random-effects model to generate mean differences.

For retrospective studies, aHRs and CIs were log-transformed [45] and pooled using a random-effects model. A *p*-value of less than 0.05 was considered statistically significant. Statistical heterogeneity was assessed using the Higgins I² index [46], calculated in CMA, with sources of heterogeneity discussed in Table 1 and expanded in the discussion. Given the limited number of studies, no funnel plot or Egger’s regression test was used to assess publication bias [47]. Study quality was assessed with the Risk of Bias (ROB) 2.0 Cochrane Tool [48] for RCTs, NHLBI Quality Assessment Tool for Before-After Studies for single arm prospective studies [49], and the Newcastle Ottawa Scale [50] for retrospective studies. No subgroup or subgroup analyses were performed

## Results

Our search yielded 10 studies [21–24,32–37] involving 13,726 patients, including 212 patients from prospective studies published between 2020 and 2024 (Table 1). One study [38] meeting the initial criteria was excluded, as it was a case series. Notably, the time periods for outcomes assessed varied significantly, ranging from 6 weeks to 6 months in prospective trials, and from 6 months to 5 years in retrospective studies. While two of the included studies were conference abstracts [24,34], the remainder were full-length, peer-reviewed articles. Risk of bias was minimal, as the studies were generally of high quality with few methodological concerns (Supplementary Table 3).

### Prospective Studies

For prospective studies assessing SGLT2I use in ascites management, the primary outcome of large-volume paracentesis (LVP) incidence was lower with SGLT2Is compared to standard of care (SoC) (RR = 0.45, CI: 0.31–0.66, *p* < 0.001, I² = 0%, *n* = 2, Figure 2A). Secondary outcomes for ascites management included a lower incidence of residual ascites (RR = 0.82, CI: 0.70–0.96, *p* = 0.012, I² = 0%, *n* = 2, Figure 2B). Compared to baseline values, there was a statistically insignificant reduction in the mean loop diuretic dose of 39.1 mg (CI: –7.77 to 83.83, *p* = 0.102, I² = 99.1%, *n* = 2, Figure 2C), but a significant reduction in mean body weight of 7.00 kg (CI: 6.04–14.44, *p* < 0.001, I² = 11.5%, *n* = 4, Figure 2D).

**Figure 2A-B.).**
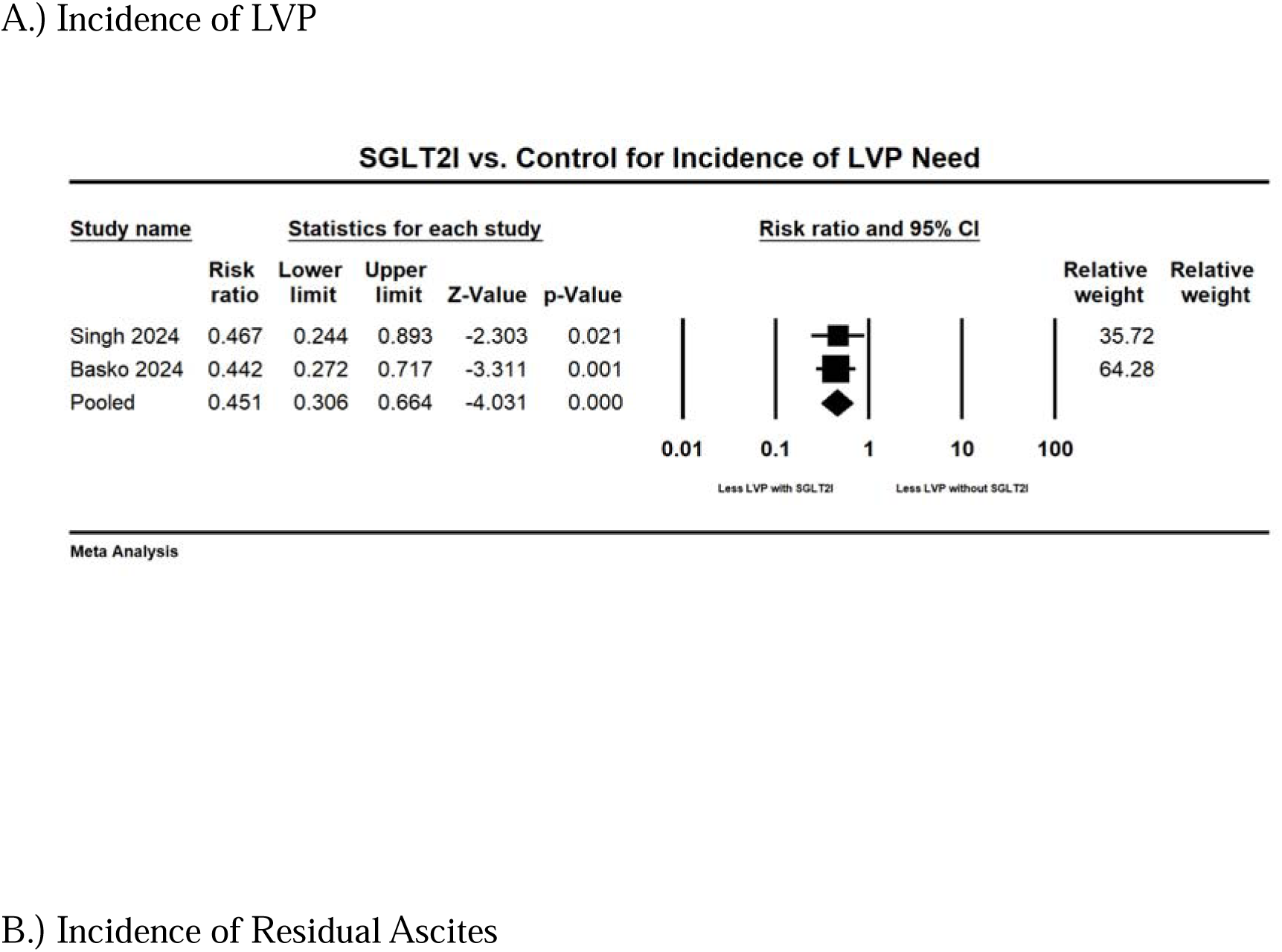
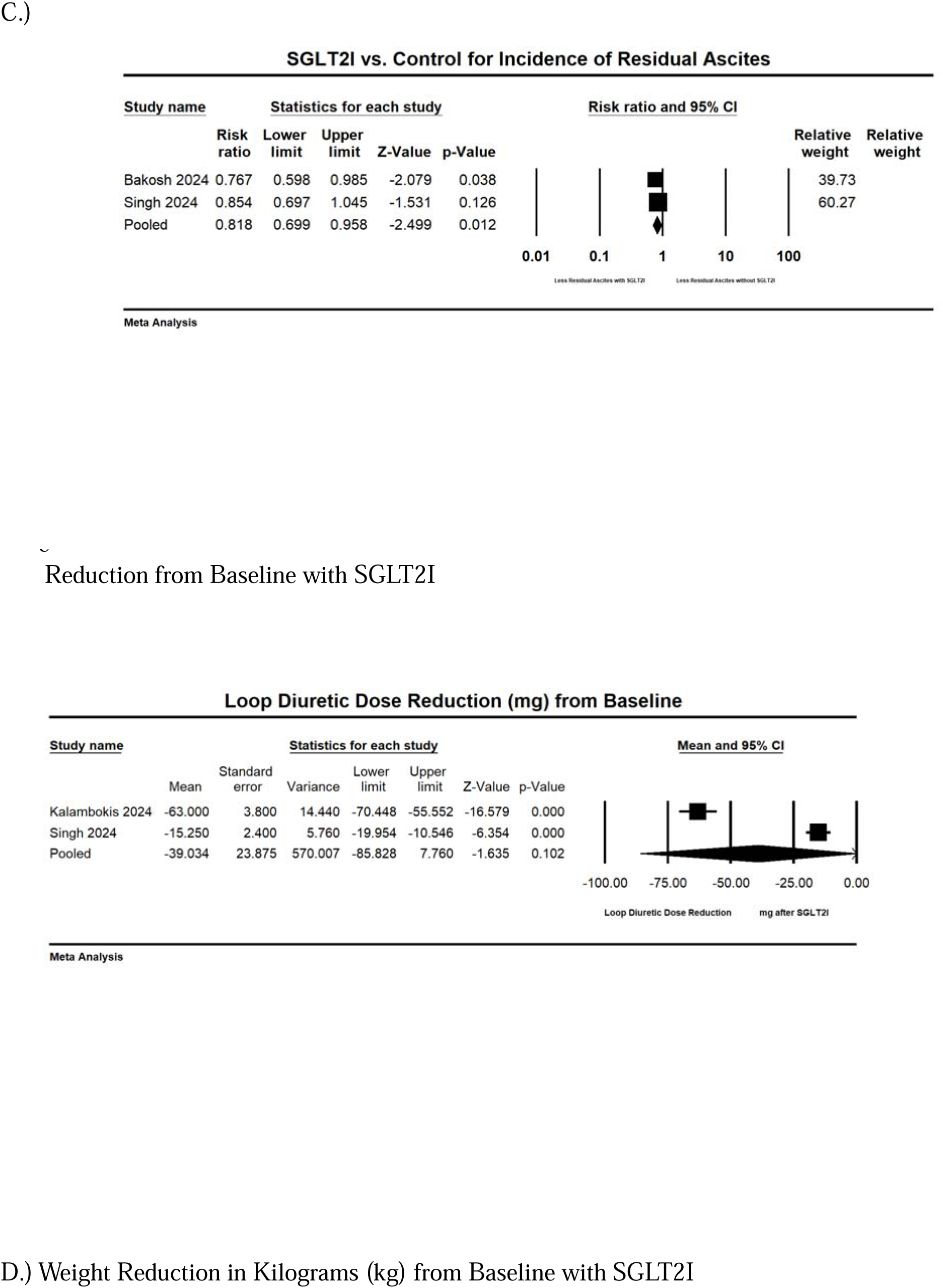
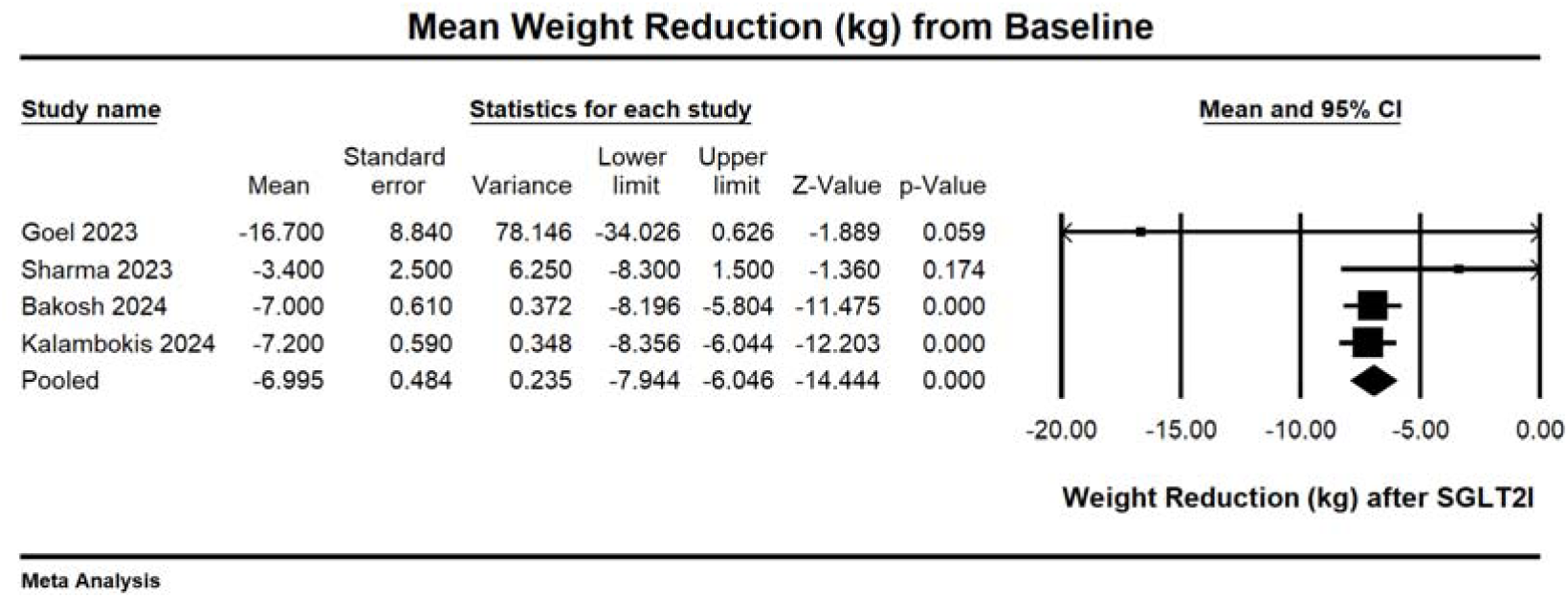
SGLT2I & Standard Diuretics vs. Standard Diuretics alone in Management in Diuretic-Resistant Ascites in Liver Cirrhosis

For adverse event-related secondary outcomes, there was no significant difference in the incidence of acute kidney injury (AKI) (RR = 1.32, CI: 0.24–7.23, *p* = 0.75, I² = 84.71%, *n* = 2, Figure 3A), new-onset hyponatraemia (RR = 2.63, CI: 0.99–6.93, *p* = 0.051, I² = 3.79%, *n* = 2, Figure 3B), hepatic encephalopathy (RR = 1.55, CI: 0.71–3.41, *p* = 0.819, I² = 0%, *n* = 2, Figure 3C), or urinary tract infections (UTIs) (RR = 2.71, CI: 0.93–7.89, *p* = 0.067, I² = 0%, Figure 3D) compared to SoC diuretics.

**Figure 3A-D.).**
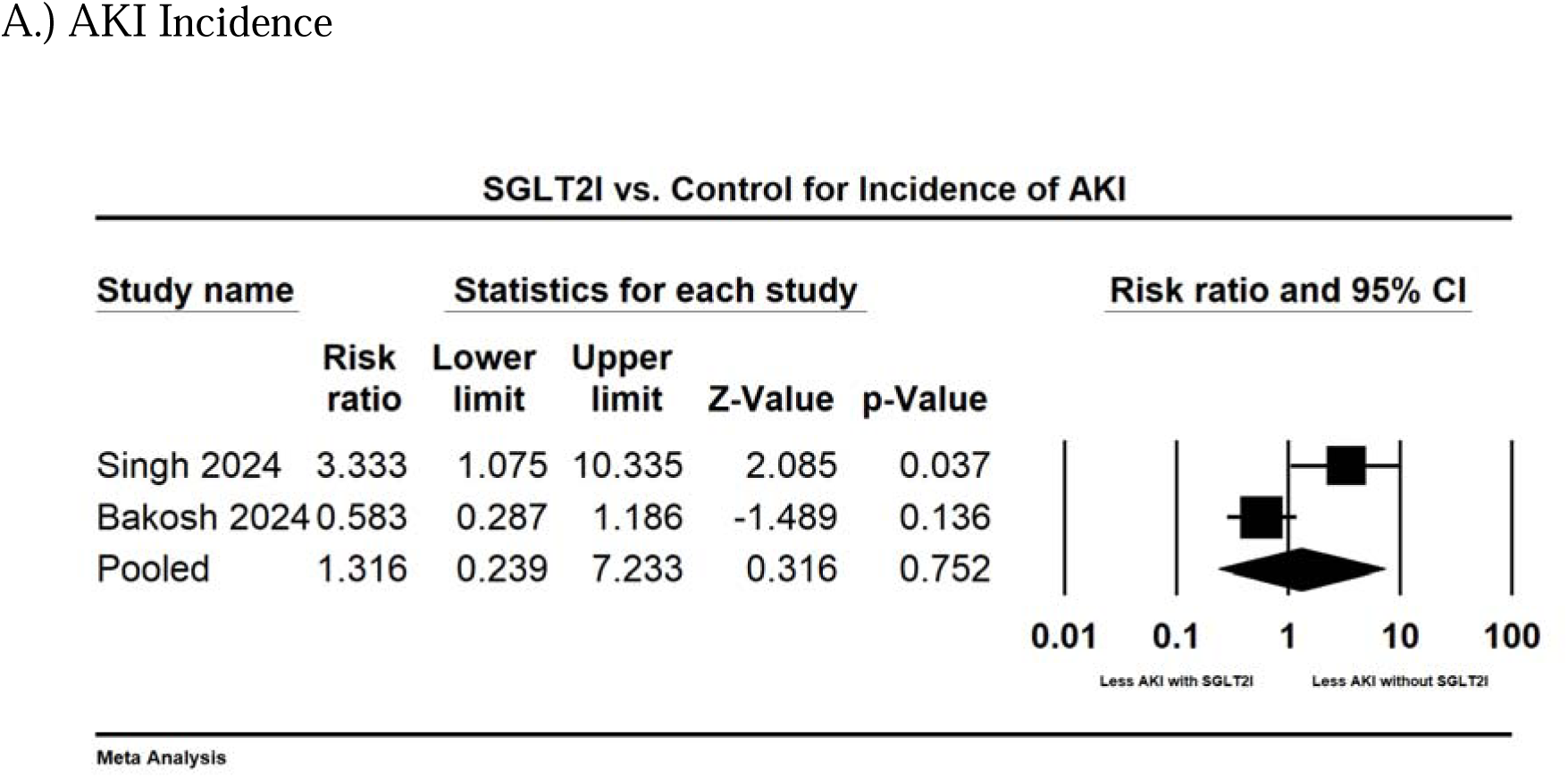
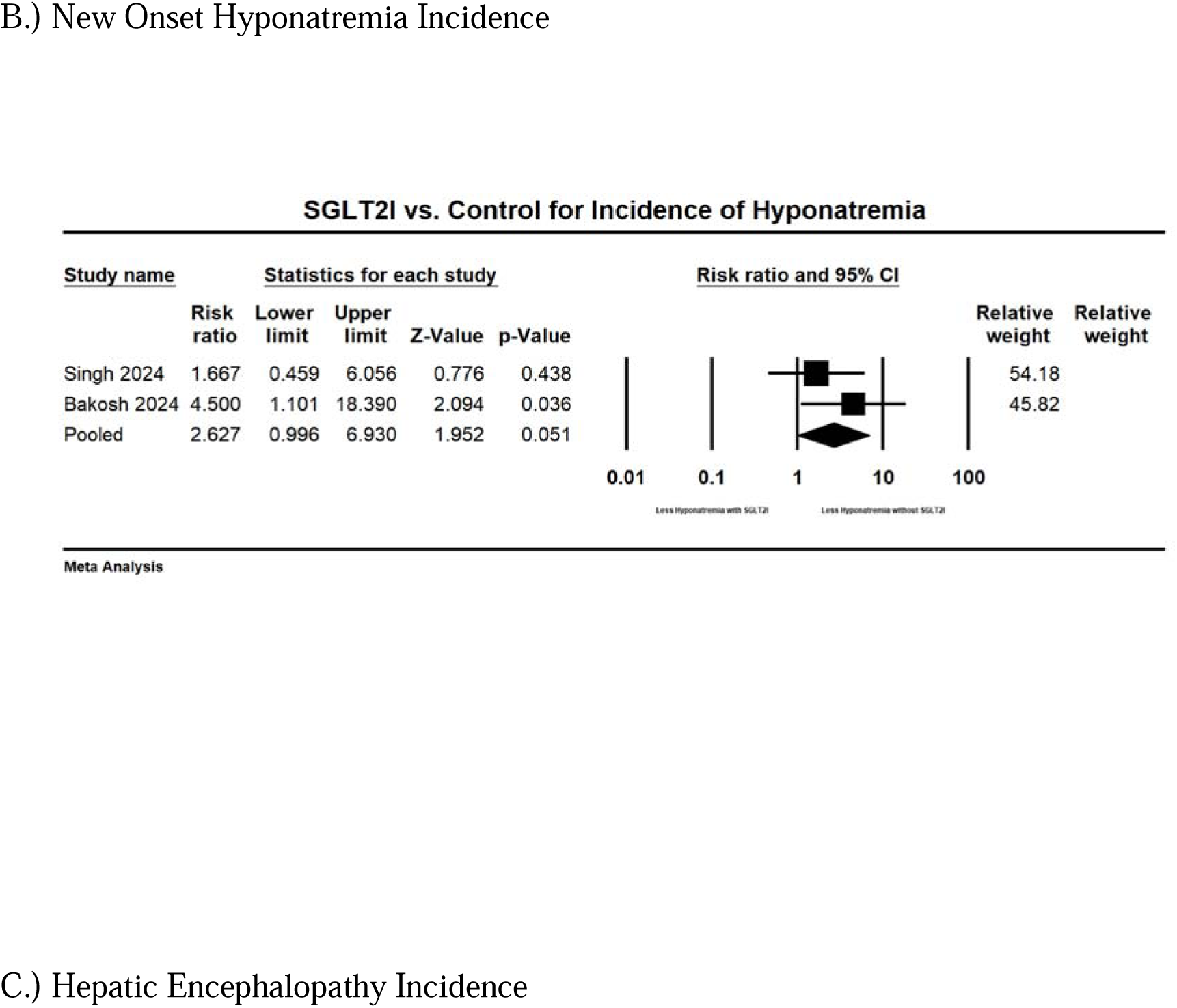
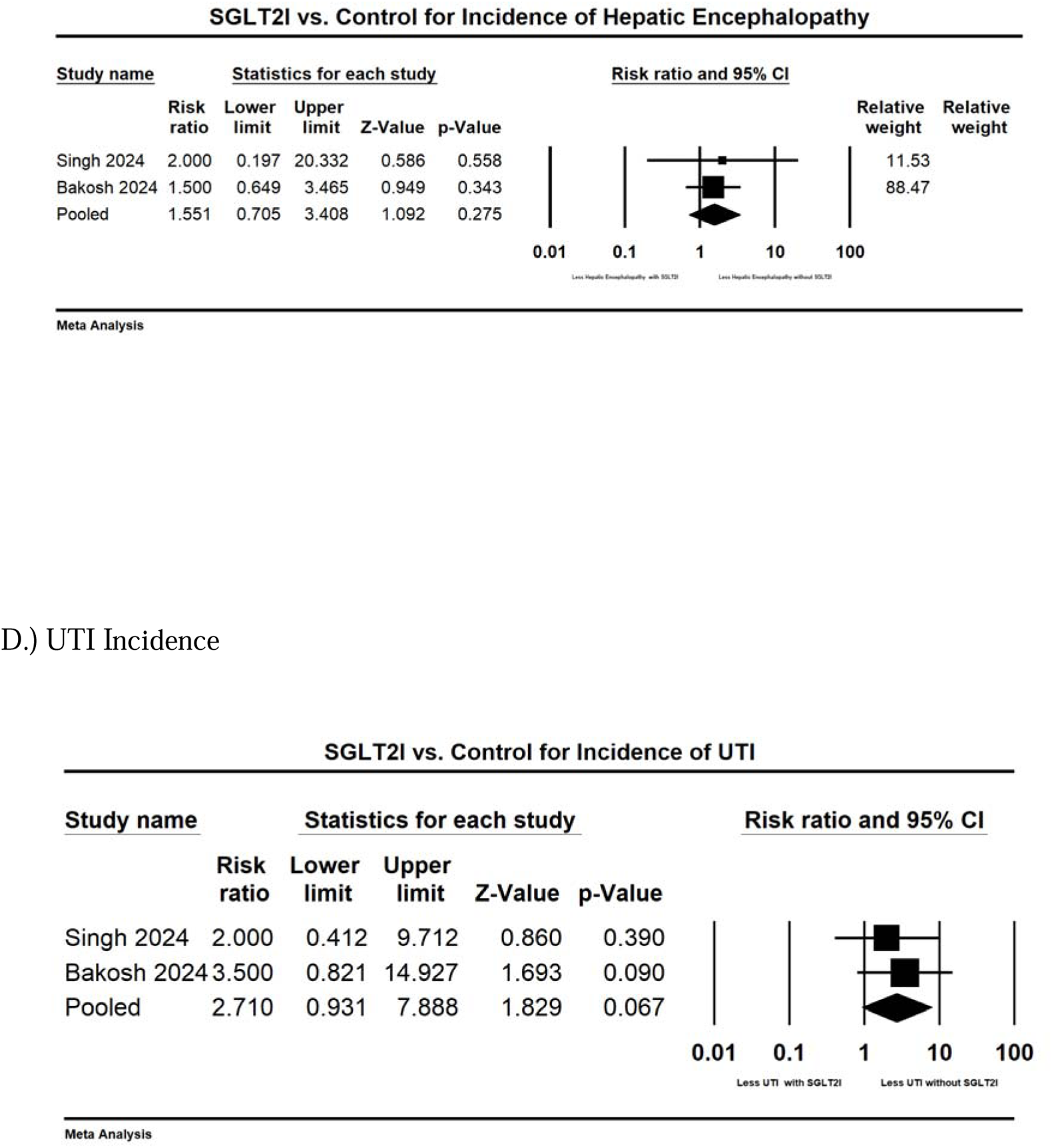
Adverse Effects SGLT2I & Standard Diuretics vs. Standard Diuretics in Diuretic-Resistant Ascites in Liver Cirrhosis

### Retrospective Studies

For retrospective studies assessing SGLT2I use in both compensated and decompensated cirrhosis, the primary outcome showed a lower mortality risk with SGLT2I use (aHR = 0.46, CI: 0.38–0.55, *p* < 0.001, I² = 26.9%, *n* = 3, Figure 4A). Secondary outcomes included a lower risk of hepatic decompensations (aHR = 0.64, CI: 0.62–0.66, *p* < 0.001, I² = 0%, *n* = 2, Figure 4B) and a lower incidence of ascites (aHR = 0.55, CI: 0.53–0.57, *p* < 0.001, I² = 0%, *n* = 2, Figure 4C).

**Figure 4A-C.).**
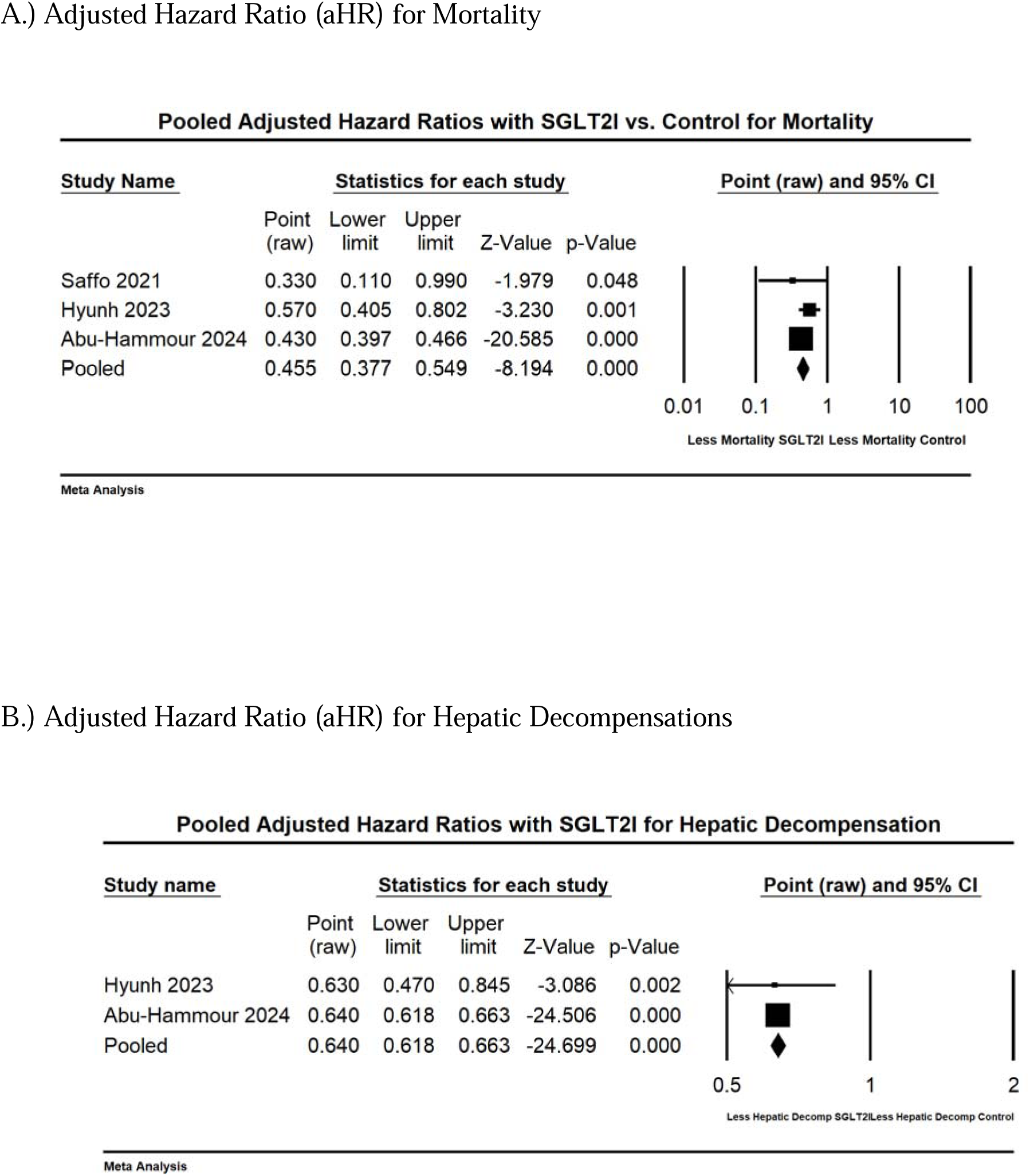
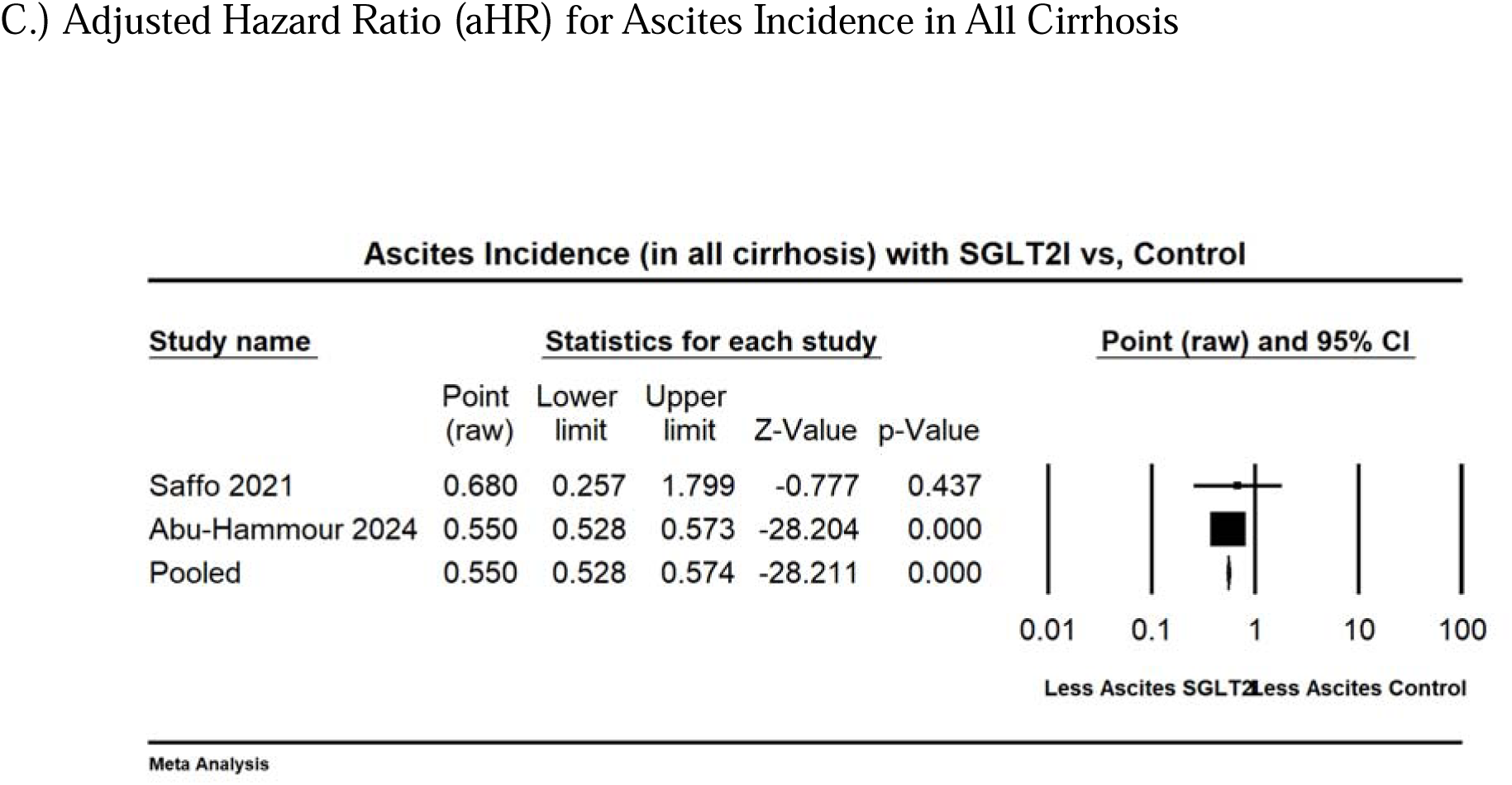
Retrospective Data on SGLT2I Exposure vs. Control

## Discussion

### Meta-Analysis Findings

Our analysis demonstrates that SGLT2Is may have utility in populations with compensated and decompensated cirrhosis while being relatively safe. In retrospective studies, SGLT2Is were associated with lower mortality, hepatic decompensation, and ascites prevalence. Prospective trials showed that adding SGLT2Is to standard diuretic therapy led to a lower need for large-volume paracentesis (LVP) with no significant differences in the incidence of acute kidney injury (AKI), new-onset hyponatraemia, hepatic encephalopathy, or urinary tract infections (UTIs). The therapy also led to clinically significant reductions in loop diuretic use and weight of approximately 40 mg and 7 kg respectively over 3 to 6 months, although the loop diuretic dose reduction did not meet statistical significance. Finally, new-onset hyponatraemia and UTIs showed a trend towards increases in the SGLT2I groups compared with control groups, despite these trends not being statistically significant.

### Clinical Implications

#### Ascites Management in Decompensated Cirrhosis

The need for large-volume paracentesis (LVP) and the prevalence of residual ascites were lower, accompanied by a reduction in weight and diuretic dose. These data were pooled from randomised controlled trials (RCTs) and single-arm trials with a low risk of bias, revealing consistent results with low calculated heterogeneity. This demonstrates a strong certainty that SGLT2Is are potent in managing diuretic-resistant ascites when used as an adjunct with spironolactone and furosemide.

In addition to weight loss reflecting ascites control, the reduction in loop diuretic dose is clinically significant, as it likely reduces side effects such as hypokalaemia [51], which correlated with one trial measuring potassium imbalances [33], showing a higher, albeit insignificant, rate of hypokalaemia in the control group. In a meta-analysis for heart failure [51], SGLT2Is in conjunction with loop diuretics led to reductions in body weight and the daily dose of loop diuretics. This aligns with our findings in liver cirrhosis, which is similarly characterised by chronic renin-angiotensin-aldosterone system (RAAS) activation [8, 30, 51].

It is important to note that most patients in these trials were diuretic-resistant [32–34, 36], meaning ascites recurred despite maximally tolerated doses of loop and mineralocorticoid receptor antagonist (MRA) diuretics. The life expectancy for patients with diuretic-resistant ascites is 6 to 12 months [52], and therefore the use of SGLT2Is could improve quality of life by reducing the necessity for hospitalisation, LVP, or diuretic titration.

It is also noteworthy that while one trial [31] measured weight loss at three months instead of six months, prospective studies [36, 37] that measured weight loss observed the greatest reduction from baseline to three months, indicating this as the most efficacious period.

Finally, many patients in our analysis receiving SGLT2Is had metabolic-associated steatotic liver disease (MASLD), which is associated with diabetes. Given that SGLT2Is exert their diuretic effect partly through glycosuria, and these drugs have been shown to be more potent diuretics in hyperglycaemic patients [53, 54], their role in ascites management may be diminished in patients with refractory ascites and cirrhosis but without diabetes. Further trials are needed to evaluate SGLT2Is in diverse aetiologies of liver cirrhosis.

### Acute Kidney Injury in Decompensated Cirrhosis

Our pooled analysis revealed no difference in the incidence of acute kidney injury (AKI) in decompensated cirrhosis. In decompensated cirrhosis, there are multiple predispositions to AKI, including splanchnic vasodilation reducing effective arterial blood volume, over-diuresis, and gastrointestinal bleeding [10, 12]. Therefore, conventional wisdom might suggest avoiding SGLT2Is to prevent precipitating renal injury.

Two randomised controlled trials (RCTs) in our analysis revealed conflicting data, effectively cancelling each other out. One trial [32] showed an insignificant increase in AKI in the control group, whereas the other [33] showed a significant increase in AKI in the SGLT2I group. Interestingly, the trial that showed increased AKI in the SGLT2I group involved patients with lower liver disease severity. The study noted that septic shock (unrelated to urinary tract infection) was overrepresented in the SGLT2I group, occurring in 12 of 13 cases of AKI [33], potentially confounding the results.

Therefore, there may even be a protective effect of SGLT2Is against AKI in decompensated cirrhosis. This is further supported by a prospective study [37] demonstrating significantly lower creatinine levels in patients treated with SGLT2Is from baseline. This effect is likely due to the same mechanism by which SGLT2Is improve renal parameters in heart failure. SGLT2Is promote afferent vasoconstriction by increasing sodium delivery to the distal tubule, thereby limiting hyperfiltration and preventing nephron damage through tubuloglomerular feedback [53].

More RCTs with larger sample sizes and homogenous aetiologies of AKI are needed to validate this conjecture based on our appraisal of the available data.

### Sodium Balance

An interesting finding of our pooled analysis was that SGLT2Is in cirrhosis trended towards an increase in new-onset hyponatraemia based on the two randomised controlled trials (RCTs). SGLT2Is promote osmotic diuresis and natriuresis through the sodium-glucose co-transporter-2 receptor; however, most existing evidence demonstrates the opposite effect. SGLT2Is typically promote more water loss than sodium loss, which would lead to possible hypernatraemia due to volume depletion, not hyponatraemia. SGLT2Is have also been cited as a potential treatment for hyponatraemia [55].

In cirrhosis, similar to heart failure, there is pre-existing free water excess leading to hyponatraemia due to chronic renin-angiotensin-aldosterone system (RAAS) activation [2, 30]. In congestive heart failure (CHF), trends have been observed where SGLT2Is increased serum sodium levels [56]. A post-hoc analysis of one trial revealed decreased serum sodium in the placebo group at two weeks, followed by an increase above baseline by eight months, and another trial showed an increase in new-onset CHF [57].

Our study’s results could be explained by several factors. First, while the incidence of new-onset hyponatraemia trended towards an increase, included prospective studies and a case series [36–38] revealed an overall increase in serum sodium from baseline. Therefore, it is possible there is an overall increase in serum sodium, but an increased incidence of new-onset hyponatraemia in some cases. Unlike heart failure patients and patients in some studies where sodium levels increased from baseline [37, 38], patients in both RCTs had diuretic-resistant ascites and were receiving high doses of diuretics, including 400 mg of spironolactone and 160 mg of furosemide. The addition of an SGLT2I in this setting may have increased the risk of hyponatraemia.

Notably, the majority of new-onset hyponatraemia cases in one trial [32] were minor, within the range of >130 mmol/L, so the clinical relevance is uncertain. More RCTs are necessary to assess the trend towards new-onset hyponatraemia with SGLT2I use in diuretic-resistant ascites, as observed in our meta-analysis.

### Hepatic Encephalopathy

As previously mentioned, SGLT2Is were shown to be associated with a trend towards hyponatraemia in the randomised controlled trials (RCTs) of our study [32, 33], as well as hypokalaemia [33], possibly due to a synergistic effect with other diuretics. Both of these electrolyte disturbances are associated with hepatic encephalopathy (HE) [13, 18], possibly through osmotic shifts [13] and renal ammonia production [58].

Additionally, hypernatraemia, dehydration, and acidosis, known side effects of SGLT2Is in the general population, are further potential triggers for HE. Exacerbation of HE was also observed in an animal study where rats with induced biliary cirrhosis were administered SGLT2Is [59]. Therefore, it is plausible to predict an increased rate of HE with SGLT2I use, particularly in patients with decompensated cirrhosis.

However, this association was not observed in our pooled analysis, which found no notable link between SGLT2I use and HE. These findings suggest that SGLT2Is are unlikely to be significant triggers of HE in decompensated cirrhosis. Nonetheless, further studies are needed to confirm this conclusion.

### Urinary Tract Infections

Cirrhotic patients are immunosuppressed due to the synthetic dysfunction of antibacterial proteins within both the innate and adaptive immune systems [16], making them susceptible to bacterial infections. Given that SGLT2Is promote glycosuria, it has been postulated that they increase the risk of bacterial urinary tract infections (UTIs) [1]. Coupled with the immunocompromise present in cirrhosis, this suggests that patients with cirrhosis would be at higher risk of developing UTIs when using SGLT2Is. However, our study only found a trend towards an increase in bacterial UTIs in decompensated cirrhosis.

This finding is consistent with data on SGLT2Is and UTIs in the general population [59], although a subgroup analysis of a meta-analysis [60] revealed a significant increase in UTIs with prolonged use beyond one year. In our study, most patients with decompensated cirrhosis were exposed to SGLT2Is for up to six months, which aligns with the life expectancy for many patients with diuretic-resistant ascites [52] and the period required to achieve a clinically relevant reduction in ascites [32, 33, 38]. Therefore, the short-term use of SGLT2Is could outweigh the risk of UTIs in diuretic-resistant ascites.

Caution should be exercised in cirrhotic patients with recent UTIs, sepsis, or other forms of immunocompromise. In one RCT [33], while the UTI rate was not higher in the SGLT2I group, there were 11 total infections in the SGLT2I group versus four in the control group, a difference that reached statistical significance. No other included study measured total infections.

Additionally, a retrospective case-control study [61] assessing the risk of developing bacteraemia in urosepsis while receiving SGLT2Is through multivariate regression found that cirrhosis was the only comorbidity significantly associated with an increased risk of bacteraemia in patients with UTI. Bacteraemia and/or sepsis can lead to acute-on-chronic liver failure [10], shock in an already vasoplegic patient [10], or seeding of the peritoneal cavity, causing spontaneous bacterial peritonitis (SBP) [62].

In patients with compensated cirrhosis, there are no head-to-head studies comparing UTI incidence with SGLT2I use versus control. Nevertheless, one might presume that the cardiovascular and reno-protective benefits of SGLT2Is would outweigh the risk of UTIs, as observed in the general population [60].

### Mortality and Hepatic Decompensation

Our analysis revealed a significant reduction in the risk of mortality and hepatic decompensation with SGLT2I use. This finding is similar to randomised controlled trials (RCTs) on the drug in congestive heart failure (CHF) [63], where SGLT2Is were noted to reduce CHF exacerbations and mortality, likely through their effects on diuresis, reduced inflammation, and improved renal function [2, 3]. These same mechanisms may be relevant in cirrhosis, where portal hypertension and fluid overload are major drivers of morbidity and mortality.

Our analysis focused more on compensated disease [22, 23] involving populations with low Model for End-Stage Liver Disease (MELD) scores. The decreased mortality and hepatic decompensation risk may stem from several mechanisms. An included single-arm study measuring haemodynamic outcomes [36] demonstrated that empagliflozin improves cardiovascular function in cirrhotic patients by decreasing systemic vascular resistance and increasing natriuresis, likely through renin-angiotensin-aldosterone system (RAAS) suppression, which has previously been documented with SGLT2I use in liver cirrhosis [30].

The consistent ability of SGLT2Is to provide reno-protective benefits, reflected by reduced albuminuria [53], may be important for cirrhotic patients, as it could delay the progression of renal dysfunction, lower the incidence of hepatorenal pathophysiology [14], and minimise sarcopenia, which is an independent predictor of mortality in cirrhosis [19]. Additionally, the diuretic effect noted in our analysis could contribute to reduced portal pressure, thereby lowering the risk of variceal development and subsequent bleeding.

Finally, as demonstrated in several RCTs [4–7], SGLT2Is may delay or improve liver fibrosis and inflammation, leading to a delay in the development of portal hypertension. Supporting this, a case series [38] found that Child-Pugh scores either decreased or remained the same over six months, and in two patients, ascites became non-detectable. Furthermore, one RCT [32] noted a three-point increase in MELD-Na scores in the control group, while the SGLT2I group’s MELD-Na scores remained near baseline.

Further long-term prospective evidence is needed to support a mortality benefit, particularly in patients with decompensated cirrhosis.

### Limitations of Meta-Analysis

Our meta-analysis has several limitations. The most significant is that the pooled analysis of prospective trials was underpowered, with only two to four effect sizes pooled per outcome.

Second, the analysis of retrospective studies is prone to selection bias, such as SGLT2Is not being prescribed to more tenuous, sicker patients. This was somewhat mitigated by propensity matching for important variables such as the Model for End-Stage Liver Disease-Sodium (MELD-Na) score [23, 24]. Additionally, retrospective studies relied on less granular data, such as International Classification of Diseases, 10th Edition (ICD-10) codes.

Third, the time periods for measuring outcomes, the types of SGLT2Is used, cirrhosis status, and the control groups to which SGLT2Is were compared varied, likely introducing heterogeneity into the analysis.

Fourth, most patients in our review had metabolic-associated steatotic liver disease (MASLD), whereas patients with other aetiologies of cirrhosis may not experience the same benefits or may have different reactions to SGLT2Is. For example, a case series documented SGLT2Is triggering diabetic ketoacidosis (DKA) in two patients with both alcoholic cirrhosis and diabetes.

Given these limitations, it is essential to carefully appraise each study’s outcomes and place our findings in the context of existing literature.

## Conclusion

Based on our systematic review and meta-analysis, the use of SGLT2Is in varying degrees of cirrhosis is associated with several benefits. These include lower mortality and a reduced hepatic decompensation rate in compensated cirrhosis, as well as a lower need for large-volume paracentesis (LVP) and loop diuretics in treatment-resistant ascites in decompensated cirrhosis. These benefits were observed without a significant increase in urinary tract infections (UTIs), new-onset hyponatraemia, or hepatic encephalopathy, although the overall risk of infection warrants further investigation.

More randomised controlled trials (RCTs) in both compensated and decompensated cirrhosis populations, with uniform outcome measures and consistent SGLT2I agents, are needed to confirm our study’s findings.

### What Is Known

- SGLT2Is confer cardiovascular and reno-protective benefits in the general population as well as those with Congestive Heart Failure (CHF)
- Recently, SGLT2I was shown to improve the metabolic profiles of patients with Metabolic-Associated Steatotic Liver Disease (MASLD), however, there is limited data on the efficacy and safety of SGTL2I in liver cirrhosis.

### What Is New

- SGLT2Is are an effective adjunct to loop diuretics and mineralocorticoids for diuretic-resistant ascites for up to 6 months, leading to clinically significant 7 kg weight loss and reduction in loop diuretic dose.
- SGLT2Is are associated with mortality benefit and decreased hepatic decompensations in cirrhosis and may have a role in slowing progression of cirrhosis.
- While SGLT2Is do not result in electrolyte derangements or increased renal injury, there may be a concern for an increased risk of overall infection while on SGLT2I which needs to be further explored, and caution should be advised in patients with recent or prior infections.

## Data Availability

All data produced in the present work are contained in the manuscript

---

**Supplementary Table 1:** Search Strategy Constructed in Embase Translated to Other Databases.

|  |
| --- |
| Keyword search terms: |
| gliflozin* OR gliflozin-derivative* OR SGLT2-inhibitor* OR sodium-dependent-glucose-cotransporter-2-inhibitor* OR sodium-glucose-co-transporter-2-inhibitor* OR sodium-glucose-transporter-2-inhibitors* OR sodium-glucose-cotransporter-2-inhibitor* OR SGLT-2-inhibit* OR SGLT2i OR SGLT2is OR SGLT2-I OR atigliflozin OR bexagliflozin OR canagliflozin OR dapagliflozin OR empagliflozin OR enavogliflozin OR ertugliflozin OR ipragliflozin OR licogliflozin OR luseogliflozin OR mizagliflozin OR remogliflozin OR sergliflozin OR sotagliflozin OR tofogliflozin |
| cirrhosis OR alcohol*-liver* OR decompensated-liver* OR hepatic-decompensat* OR liver-decompensat* |
| Embase.com Searches (with Emtree Search headings). Do not use subheadings unless you intend to exclude Conference Abstracts<br>'sodium glucose cotransporter 2 inhibitor'/syn<br>'liver cirrhosis'/syn NOT (('fatty liver'/exp/mj OR NAFLD:ti OR MAFLD:ti OR MASLD:ti OR fatty-liver:ti) NOT ('liver cirrhosis'/exp/mj NOT cirrhos*:ti))<br>NOT ([animals]/lim NOT [humans]/lim)<br>NOT ('conference review'/it OR 'editorial'/it OR 'letter'/it OR 'note'/it OR 'review'/it OR 'short survey'/it OR 'tombstone'/it OR 'case report'/de OR 'meta analysis'/de OR 'meta analysis topic'/de OR 'systematic review'/de OR 'systematic review topic'/de) |
| [MEDLINE in PubMed](http://MEDLINE.in.PubMed) (with MeSH headings). Use OVID MEDLINE if you are using proximity operators.<br>"Liver Cirrhosis"[Mesh] NOT (("Fatty Liver"[Majr] OR NAFLD[ti] OR MAFLD[ti] OR MASLD[ti] OR fatty-liver[ti]) NOT ("Liver Cirrhosis"[Majr] OR cirrhos*[ti]))<br>"Sodium-Glucose Transporter 2 Inhibitors"[Mesh] OR "Sodium-Glucose Transporter 2 Inhibitors" [Pharmacological Action]<br>NOT ("animals"[mesh] NOT "humans"[mesh])<br>NOT ("case reports"[Publication Type] OR "comment"[Publication Type] OR "editorial"[Publication Type] OR "guideline"[Publication Type] OR "introductory journal article"[Publication Type] OR "meta analysis"[Publication Type] OR "news"[Publication Type] OR "retracted publication"[Publication Type] OR "review"[Publication Type] OR "systematic review"[Publication Type]) |
| [Cochrane Central Register of Controlled Trials](http://Cochrane.Central.Register.of.Controlled.Trials). Use MEDLINE search but "Term"[MeSH] > [mh "Term"]. Limit to "Trials", so results are automatically limited to Controlled Trial studies and Trial Registries, so no need for publication type limits. |
| Keywords for [Global Index Medicus](http://Global.Index.Medicus) – remove truncation from keywords. Use "" around phrases. |

**Supplementary Table 2:**

| Database | Results | Platform |
| --- | --- | --- |
| Embase | 167 | Embase.com (Elsevier) |
| MEDLINE | 58 | PubMed (NCBI, National Libraries of Medicine) |
| Cochrane Central Register of Controlled Trials | 36 | Cochrane Library (Wiley) |
| Web of Science Core Collection | 95 | Web of Science (Clarivate) |
| Total | 356 |  |
| with duplicates removed | 255 |  |

**Supplementary Table 3A:** Quality Assessment of Included Studies Using Cochrane RoB 2.0 Tool, NIH, and Newcastle-Ottawa Scale for Retrospective Studies.

| ROB 2.0 | Randomization | Comments | Intended Intervention | Comments | Outcome Data | Comments | Measurement of Outcomes | Comments | Reported Results | Comments | Overall | Notes |
| --- | --- | --- | --- | --- | --- | --- | --- | --- | --- | --- | --- | --- |
| Bokosh 2024 | Low | Block randomization, also no difference in baseline characteristics | Low | Assessor -blinded, no missed doses | Low | Primary outcome reflects aim | Low Risk | Assessor-blind, objective data | Low Risk | No outcome data omitted | Low Risk | Block randomization (SNOSE), No difference in baseline characteristics, assessor-blinded, outcomes reflect aim, no outcome data omitted. |
| Singh 2024 | Low | Double-blind, randomized, placebo-controlled trial. Randomization was done using a computer-generated random number trial | Low | Allocation concealment was done using the serially numbered opaque sealed envelopes (SNOSE) method | Low | Primary outcome reflects aim | Low Risk | Clinicians were blinded to group assignments. Key outcomes were objective measures. Patients were monitored frequently. | Unclear risk | 12 of the 40 patients (30%) died before trial completed | Low Risk | Double blind, randomized, placebo-controlled. Study followed timeline submitted to clinicaltrials.gov. Reported both positive and negative findings |

**Supplementary Table 3B:**
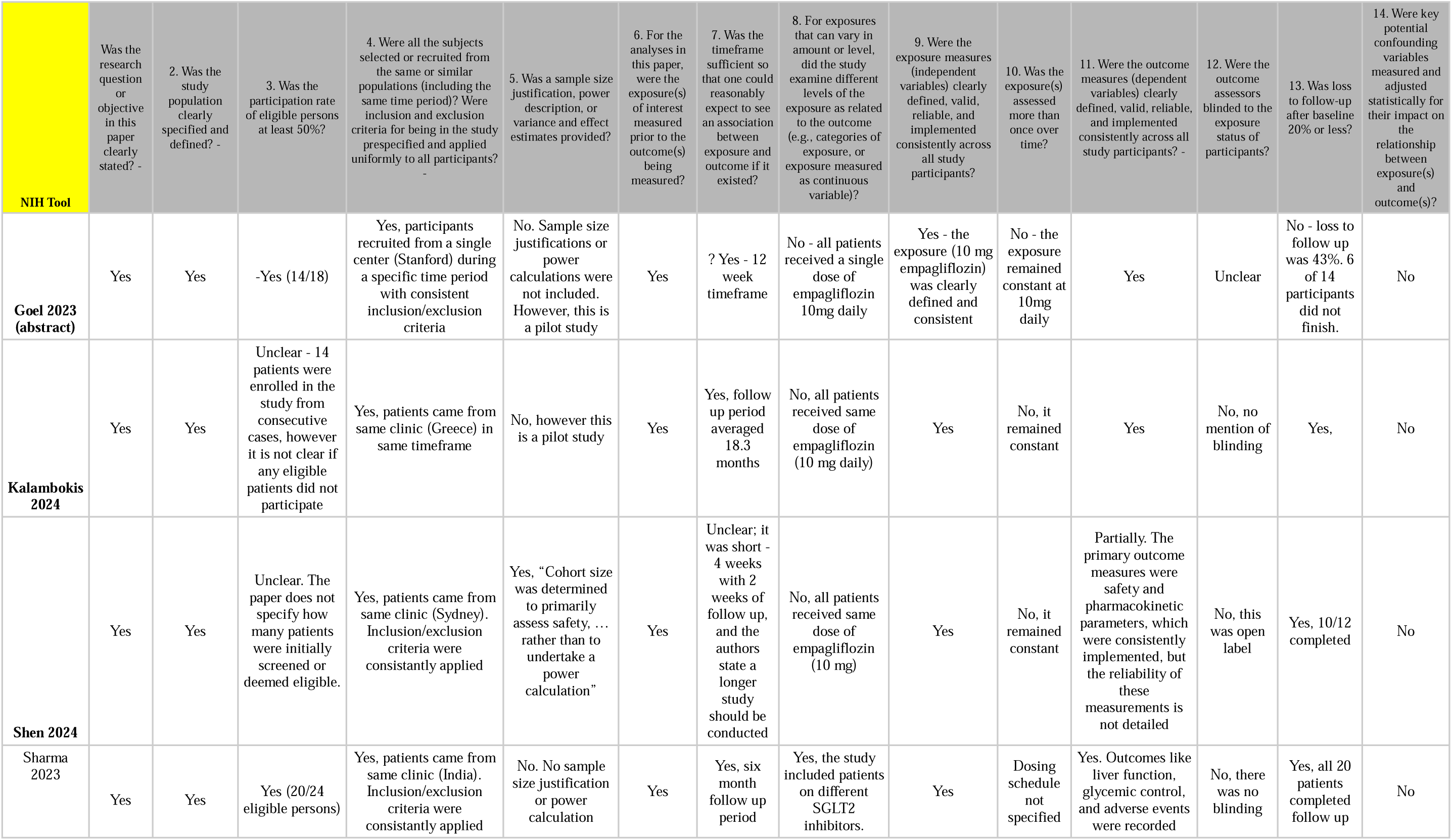
Quality Assessment of Included Studies using National Institutes of Health (NIH) Quality Assessment Tool for Observational Studies and Cross-Sectional Studies.

**Supplementary Table 3C:**
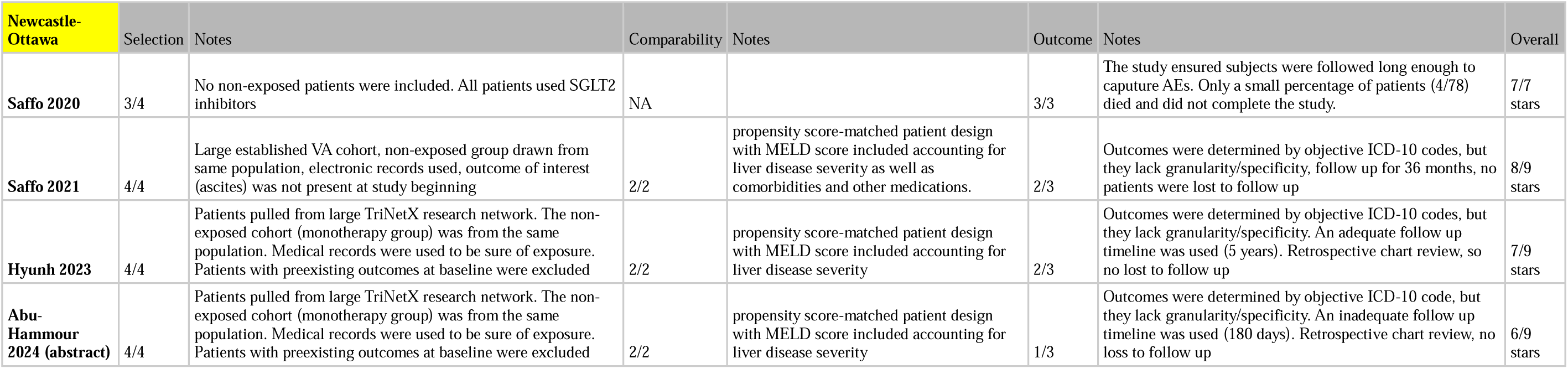

| Newcastle-Ottawa | Selection | Notes | Comparability | Notes | Outcome | Notes | Overall |
| --- | --- | --- | --- | --- | --- | --- | --- |
| **Saffo 2020** | 3/4 | No non-exposed patients were included. All patients used SGLT2 inhibitors | NA |  | 3/3 | The study ensured subjects were followed long enough to capture AEs. Only a small percentage of patients (4/78) died and did not complete the study. | 7/7 stars |
| **Saffo 2021** | 4/4 | Large established VA cohort, non-exposed group drawn from same population, electronic records used, outcome of interest (ascites) was not present at study beginning | 2/2 | propensity score-matched patient design with MELD score included accounting for liver disease severity as well as comorbidities and other medications. | 2/3 | Outcomes were determined by objective ICD-10 codes, but they lack granularity/specificity, follow up for 36 months, no patients were lost to follow up | 8/9 stars |
| **Hyunh 2023** | 4/4 | Patients pulled from large TriNetX research network. The non-exposed cohort (monotherapy group) was from the same population. Medical records were used to be sure of exposure. Patients with preexisting outcomes at baseline were excluded | 2/2 | propensity score-matched patient design with MELD score included accounting for liver disease severity | 2/3 | Outcomes were determined by objective ICD-10 codes, but they lack granularity/specificity. An adequate follow up timeline was used (5 years). Retrospective chart review, so no lost to follow up | 7/9 stars |
| **Abu-Hammour 2024 (abstract)** | 4/4 | Patients pulled from large TriNetX research network. The non-exposed cohort (monotherapy group) was from the same population. Medical records were used to be sure of exposure. Patients with preexisting outcomes at baseline were excluded | 2/2 | propensity score-matched patient design with MELD score included accounting for liver disease severity | 1/3 | Outcomes were determined by objective ICD-10 code, but they lack granularity/specificity. An inadequate follow up timeline was used (180 days). Retrospective chart review, no loss to follow up | 6/9 stars |

**Supplementary Table 4:**

| Section and Topic | Item # | Checklist item | Location where item is reported |
| --- | --- | --- | --- |
| **TITLE** |  |  |  |
| Title | 1 | Identify the report as a systematic review. | 1-2 |
| **ABSTRACT** |  |  |  |
| Abstract | 2 | See the PRISMA 2020 for Abstracts checklist. | 3 |
| **INTRODUCTION** |  |  |  |
| Rationale | 3 | Describe the rationale for the review in the context of existing knowledge. | 4-5 |
| Objectives | 4 | Provide an explicit statement of the objective(s) or question(s) the review addresses. | 4-5 |
| **METHODS** |  |  |  |
| Eligibility criteria | 5 | Specify the inclusion and exclusion criteria for the review and how studies were grouped for the syntheses. | 6 |
| Information sources | 6 | Specify all databases, registers, websites, organisations, reference lists and other sources searched or consulted to identify studies. Specify the date when each source was last searched or consulted. | 6 |
| Search strategy | 7 | Present the full search strategies for all databases, registers and websites, including any filters and limits used. | 6, Supplementary Table 1-2 |
| Selection process | 8 | Specify the methods used to decide whether a study met the inclusion criteria of the review, including how many reviewers screened each record and each report retrieved, whether they worked independently, and if applicable, details of automation tools used in the process. | 6 |
| Data collection process | 9 | Specify the methods used to collect data from reports, including how many reviewers collected data from each report, whether they worked independently, any processes for obtaining or confirming data from study investigators, and if applicable, details of automation tools used in the process. | 7 |
| Data items | 10a | List and define all outcomes for which data were sought. Specify whether all results that were compatible with each outcome domain in each study were sought (e.g. for all measures, time points, analyses), and if not, the methods used to decide which results to collect. | 7 |
|  | 10b | List and define all other variables for which data were sought (e.g. participant and intervention characteristics, funding sources). Describe any assumptions made about any missing or unclear information. | NA |
| Study risk of bias assessment | 11 | Specify the methods used to assess risk of bias in the included studies, including details of the tool(s) used, how many reviewers assessed each study and whether they worked independently, and if applicable, details of automation tools used in the process. | 8-9 |
| Effect measures | 12 | Specify for each outcome the effect measure(s) (e.g. risk ratio, mean difference) used in the synthesis or presentation of results. | 8-9 |
| Synthesis methods | 13a | Describe the processes used to decide which studies were eligible for each synthesis (e.g. tabulating the study intervention characteristics and comparing against the planned groups for each synthesis (item #5)). | 7-8 |
|  | 13b | Describe any methods required to prepare the data for presentation or synthesis, such as handling of missing summary statistics, or data conversions. | 7-8 |
|  | 13c | Describe any methods used to tabulate or visually display results of individual studies and syntheses. | 8-9 |
|  | 13d | Describe any methods used to synthesize results and provide a rationale for the choice(s). If meta-analysis was performed, describe the model(s), method(s) to identify the presence and extent of statistical heterogeneity, and software package(s) used. | 8-9 |
|  | 13e | Describe any methods used to explore possible causes of heterogeneity among study results (e.g. subgroup analysis, meta-regression). | 8-9 |
|  | 13f | Describe any sensitivity analyses conducted to assess robustness of the synthesized results. | NA |
| Reporting bias assessment | 14 | Describe any methods used to assess risk of bias due to missing results in a synthesis (arising from reporting biases). | 9 |
| Certainty assessment | 15 | Describe any methods used to assess certainty (or confidence) in the body of evidence for an outcome. | 8-9 |
| **RESULTS** |  |  |  |
| Study selection | 16a | Describe the results of the search and selection process, from the number of records identified in the search to the number of studies included | 9 |
|  |  | in the review, ideally using a flow diagram. | Figure 1 |
|  | 16b | Cite studies that might appear to meet the inclusion criteria, but which were excluded, and explain why they were excluded. | 9 |
| Study characteristics | 17 | Cite each included study and present its characteristics. | 9<br>Table 1 |
| Risk of bias in studies | 18 | Present assessments of risk of bias for each included study. | 10<br>Supplementary Table 3 |
| Results of individual studies | 19 | For all outcomes, present, for each study: (a) summary statistics for each group (where appropriate) and (b) an effect estimate and its precision (e.g. confidence/credible interval), ideally using structured tables or plots. | 9-10 |
| Results of syntheses | 20a | For each synthesis, briefly summarise the characteristics and risk of bias among contributing studies. | Table 1<br>Supplementary Table 3 |
|  | 20b | Present results of all statistical syntheses conducted. If meta-analysis was done, present for each the summary estimate and its precision (e.g. confidence/credible interval) and measures of statistical heterogeneity. If comparing groups, describe the direction of the effect. | 9-10<br>Figure 2-4 |
|  | 20c | Present results of all investigations of possible causes of heterogeneity among study results. | 9-10<br>Table 1 |
|  | 20d | Present results of all sensitivity analyses conducted to assess the robustness of the synthesized results. | NA |
| Reporting biases | 21 | Present assessments of risk of bias due to missing results (arising from reporting biases) for each synthesis assessed. | NA given limited studies |
| Certainty of evidence | 22 | Present assessments of certainty (or confidence) in the body of evidence for each outcome assessed. | 9-16 |
| **DISCUSSION** |  |  |  |
| Discussion | 23a | Provide a general interpretation of the results in the context of other evidence. | 10 |
|  | 23b | Discuss any limitations of the evidence included in the review. | 17-18 |
|  | 23c | Discuss any limitations of the review processes used. | 17-18 |
|  | 23d | Discuss implications of the results for practice, policy, and future research. | 1; 11-18 |
| **OTHER INFORMATION** |  |  |  |
| Registration and protocol | 24a | Provide registration information for the review, including register name and registration number, or state that the review was not registered. | 1 |
|  | 24b | Indicate where the review protocol can be accessed, or state that a protocol was not prepared. | 1 |
|  | 24c | Describe and explain any amendments to information provided at registration or in the protocol. | 1 |
| Support | 25 | Describe sources of financial or non-financial support for the review, and the role of the funders or sponsors in the review. | 1 |
| Competing interests | 26 | Declare any competing interests of review authors. | 1 |
| Availability of data, code and other materials | 27 | Report which of the following are publicly available and where they can be found: template data collection forms; data extracted from included studies; data used for all analyses; analytic code; any other materials used in the review. | 1 |

